# Anxiety disorders and asthma among adolescents in urban Uganda: the role of early life exposures

**DOI:** 10.1101/2020.10.08.20209478

**Authors:** Harriet Mpairwe, Richard Stephen Mpango, Wilber Sembajjwe, Emily L Webb, Alison M Elliott, Neil Pearce, Eugene Kinyanda

## Abstract

**Background:** The reasons for the association between anxiety disorders and asthma are not fully established, and data from Africa is sparse. We investigated whether the association between anxiety disorders and asthma among adolescents may be partly related to shared exposures in early life.

**Methods:** We conducted a case-control study among adolescents (12-17 years) with and without asthma in Wakiso District, an urban area in Uganda. Anxiety disorders were diagnosed by the Youth Inventory-4R (YI-4R), a Diagnostic and Statistical Manual of Mental Disorders, Fifth Edition (DSM-5) referenced instrument. For this report, we focus on generalized anxiety disorder (GAD), panic disorder and social anxiety disorder. Asthma was doctor-diagnosed by study clinicians. We used questionnaires to collect data on early life exposures. The data were analysed using multiple logistic regression models.

**Results:** We enrolled 162 adolescents. Adolescents with asthma were more likely to have any of three anxiety disorders (44.6%) than adolescents without asthma (21.4%) [adjusted odds ratio (AOR) 2.68, 95% confidence interval (CI) 1.30-5.53, p-value=0.007]. The association was strong for GAD (AOR 4.49, 95% CI 1.48-13.56) and panic disorder (AOR 5.43, 95% CI 2.11-14.02), but not for social anxiety disorder (1.46, 95% CI 0.63-3.37). The early life risk factors associated with anxiety disorders among adolescents were similar to asthma risk factors previously published, including urban residence at birth [AOR 3.42 (1.29-9.09)] and during most of the first five years of life [AOR 2.87 (1.07-7.66)], father’s tertiary education [AOR 2.09 (1.00-4.37)], and adolescent’s history of other allergy-related diseases [AOR 4.64 (1.66-13.00)].

**Conclusion:** We confirm a positive association between anxiety disorders and asthma among adolescents in urban Uganda. The early life risk factors associated with anxiety disorders among adolescents were similar to those for asthma in the same age-group, suggesting shared underlying causes.

## Introduction

Anxiety disorders are among the first psychiatric disorders to emerge in childhood and adolescence^1, 2^, and are the sixth leading cause of illness and disability among adolescents aged 10–14 years and the ninth for adolescents aged 15–19 years globally^1^. Anxiety disorders are common, with global estimates for lifetime prevalence of 9.9-16.7% (inter-quartile range)^3^. Anxiety disorders, as classified by the Diagnostic and Statistical Manual of Mental Disorders-Fifth Edition (DSM-5), include generalized anxiety disorder, social anxiety disorder, separation anxiety disorder, specific phobia, and panic disorder^2^. The causes of anxiety disorders are not fully known but environmental, developmental, and biological risk factors in early life have been identified^2^. Adolescents are at greater risk of anxiety disorders if they have chronic illnesses, such as asthma^1^.

Asthma is the most common non-communicable disease among children, and is estimated to affect 235 million people globally, yet we still do not fully understand the causes^4^. Several early-life risk factors for asthma have been suggested including parental asthma, prenatal environmental tobacco smoke exposure and prematurity^5^. It has been observed that in Africa and other low- and middle-income countries (LMICs), the prevalence of asthma is higher in urban than rural areas^6-9^. Our study in Uganda found that the risk of asthma depended on area of residence at birth and in the first five years: lowest for rural-born children, with risk doubling among town-born children and trebling among city-born children^10^. This and other studies highlight the importance of environmental exposures in early life for the development of asthma in later childhood^5, 11^.

Asthma and anxiety disorders are positively associated among children, adolescents and adults^12, 13^. The reasons for this have not been fully resolved, resulting in an on-gong *chicken and egg* debate on which comes first, with some authors suggesting a bidirectional relationship^14, 15^. In addition, a third alternative to possibly explain the positive association between anxiety disorders and asthma has been suggested as shared liability, through shared environmental exposures^16^. There is a scarcity of data on anxiety disorders and asthma among adolescents in sub-Saharan Africa. We investigated the associations between anxiety disorders and asthma among adolescents in urban Uganda, and the associations between reported exposures in early life and anxiety disorders in adolescents.

## Methods

### Study design and enrolment procedures

This study was nested within a larger asthma case-control study that has previously been reported^10^. Briefly, the parent case-control study enrolled schoolchildren, 5-17 years, with and without asthma, from schools in Wakiso District, an urban area in central Uganda. The study was conducted from May 2015 to July 2017, and aimed to recruit all schoolchildren with asthma and a random sample of two non-asthma controls for each case. For this nested study, we collected data on anxiety disorders from adolescents (12-17 years) who enrolled into the parent case-control study between the months of March and August 2016. This data collection involved answering additional questions to diagnose anxiety disorders (details below), administered by two experienced psychiatric clinical officers (PCOs), supervised by a clinical psychologist and psychiatrist. We defined adolescents as children 12-17 years of age (World Health Organisation definition is 10-19 years^17^); 12 years was the required minimum age for the key diagnostic tool for anxiety disorders (below) and 17 years was the upper age limit for the parent case-control study^10^.

### Definition of anxiety disorders

We used the Diagnostic and Statistical Manual of Mental Disorders, Fifth Edition (DSM-5) referenced instrument, the Youth Inventory-4R (YI-4R), to diagnose anxiety disorders^18^. The Y1-4R is a youth-reported scale that provides invaluable insight into how youth perceive their problems. The symptom/sign and impairment items, and how these were scored to generate the diagnostic categories, as well as the reliability and validity of this scale among young people in Uganda has been reported previously^19^. For this paper, we focused on three anxiety disorders whose onset typically occurs during adolescence including generalized anxiety disorder (GAD), panic disorder and social anxiety disorder^2^. On average, assessment with the YI-4R scale lasted for approximately 45 minutes. Any adolescents who required psychiatric management received initial attention from the psychiatric clinical officers and were referred for further management, appropriately.

### Asthma diagnosis and control

Adolescents were screened for asthma using the International Study of Asthma and Allergies in Childhood (ISAAC) questionnaire^20^. Adolescents with a history of wheezing in the last 12 months underwent a clinical assessment by study clinicians which involved detailed medical and treatment history to diagnose asthma^10^. In addition, we assessed asthma control in the last four weeks using the childhood Asthma Control Test (cACT)^21^ for 12-year old adolescents and the Asthma Control Test (ACT)^22^ for those aged 13 years and above. Adolescents with no history of wheezing or any asthma symptoms were enrolled as non-asthma controls^10^.

### Additional data collected

We used questionnaires to collect data from parents or guardians on potential early life risk factors such as area of residence at the time of birth (proxy for residence in pregnancy), main area of residence at 0-5 years of age for the adolescent, parental education and history of allergy. We also collected data from adolescents on reported triggers for asthma symptoms. We performed skin prick tests (SPT) for allergic sensitisation using seven crude extracts (*Dermatophagoides* mix of *D. farinae* and *D. pteronyssminus, Blomia tropicalis, Blattella germanica*, peanut, cat, pollen mix of weeds, mould mix of *Aspergillus* species; ALK Abello, Hoersholm, Denmark), as well as a saline negative control and histamine positive control^23^. We used the standard procedures for the SPT procedure as previously described^10, 23^.

### Ethical considerations

Adolescents provided written informed assent and their parents or guardians provided written informed consent. The consenting process was conducted in either English or Luganda (the main language in the study area). The study was conducted according to Good Clinical Practice guidelines and obtained ethical approval from two independent bodies: Uganda Virus Research Institute Research and Ethics Committee (reference number GC/127/14109/481) and Uganda National Council for Science and Technology (reference number HS 1707).

### Statistical methods

Data were collected on pre-coded paper questionnaires and double entered in OpenClinica open source software version 3.1.4 (OpenClinica LLC and collaborators, Waltham, MA, USA). Data were analysed in STATA version 15 (StataCorp, Texas, USA).

We used multivariable logistic regression to estimate odds ratios and 95% confidence intervals. We adjusted for confounders such as age and sex determined a priori; father’s highest education status, as suggested in literature; and for adolescents’ residence at birth, where appropriate. Multiple linear regression models were fitted for asthma control test scores, and adjusted for confounders as above. For the purpose of this analysis, the outcomes in the regression models varied for the different analyses: for the association between anxiety disorders and asthma and with asthma control, asthma and asthma control scores were considered the outcome; for the association between early life risk factors and anxiety disorders, anxiety disorders were considered the outcome.

## Results

The details of the numbers of participants enrolled in the parent case-control study have been described elsewhere^10^. Briefly, 1,702 schoolchildren were enrolled, 562 with and 1,140 without asthma, from 55 schools in Wakiso. This nested study on anxiety disorders enrolled 162 adolescents, 73 with and 89 without asthma; the mean age was 14.3 years (range 12 to 17) and 111 (68.5%) of these were girls (girls were 55.9% in the parent case-control study).

### Anxiety disorders and asthma

The proportion of children reporting anxiety disorders was different for adolescents with and without asthma; the overall prevalence of any of the three anxiety disorders was higher (46.6%) among adolescents with asthma than among adolescents without asthma (21.4%); (adjusted odds ratio (AOR) 3.05, 95% confidence interval (CI) 1.50-6.21), **Table 1**). This difference was particularly strong for general anxiety disorder (AOR 5.16, 95% CI 1.71-15.55) and panic disorder (AOR 5.92, 95% CI 2.31-15.19), but not for social anxiety disorder (1.68, 95% CI 0.74-3.84, **Table 1**).

**Table 1:** Anxiety disorders among adolescents with and without asthma in Uganda (N=162)

| Anxiety disorders | Asthma cases<br>N=73 | Non-asthma controls<br>N=89 | Crude OR (95% CI) | Adj. OR<sup>‡</sup> (95% CI) | P-value |
| --- | --- | --- | --- | --- | --- |
| **Any of 3 anxiety disorders** |  |  |  |  |  |
| No (109) | 39 (53.4) | 70 (78.6) | 1 | 1 |  |
| Yes (53) | 34 (46.6) | 19 (21.4) | 3.21 (1.62-6.37) | 3.05 (1.50-6.21) | 0.007 |
| **General anxiety disorder** |  |  |  |  |  |
| No (141) | 57 (78.1) | 84 (94.4) | 1 | 1 |  |
| Yes (21) | 16 (21.9) | 5 (5.6) | 4.71 (1.63-13.60) | 5.16 (1.71-15.55) | 0.008 |
| **Panic disorder** |  |  |  |  |  |
| No (130) | 49 (67.1) | 81 (91.0) | 1 | 1 |  |
| Yes (32) | 24 (32.9) | 8 (9.0) | 4.96 (2.07-11.90) | 5.92 (2.31-15.19) | <0.0001 |
| **Social anxiety disorder** |  |  |  |  |  |
| No (131) | 55 (75.3) | 76 (85.4) | 1 | 1 |  |
| Yes (31) | 18 (24.7) | 13 (14.6) | 1.91 (0.86-4.23) | 1.68 (0.74-3.84) | 0.38 |

N=number; CI=confidence interval; Adj. OR=adjusted odds ratio. 2<sup>nd</sup> and 3<sup>rd</sup> columns show numbers (percentages). <sup>‡</sup>Adjusted for child's age, sex and father's highest education level attained.

### Anxiety disorders and asthma control

Asthma control test scores were generated from the Asthma Control Test^24^. Asthma control was poorer among adolescents with any of the three anxiety disorders, as depicted by the lower asthma control test scores (adjusted mean difference −2.66, 95% CI −5.01, −0.30), **Table 2**. A similar pattern was observed for all the individual anxiety disorders, but the differences were not statistically significant, **Table 2**.

**Table 2:** Anxiety disorders and asthma control among adolescents with in Uganda (N=72)

| Anxiety disorder | Asthma control test scores<sup>a</sup> |  |  |  |
| --- | --- | --- | --- | --- |
|  | Univariate analysis |  | Multivariate analysis<sup>‡</sup> |  |
|  | Mean (SD) | p-value | Adjusted mean difference (95% CI) | p-value |
| Any of 3 anxiety disorders |  |  |  |  |
| No (38) | 21.24 (5.17) |  | ref | 0.03 |
| Yes (34) | 18.26 (4.93) | 0.01 | -2.66 (-5.01, -0.30)<sup>∞</sup> |  |
| Generalised anxiety disorder |  |  |  |  |
| No (56) | 20.48 (5.40) |  | ref | 0.11 |
| Yes (16) | 17.56 (3.98) | 0.05 | -2.34 (-5.20, 0.52) |  |
| Panic Disorder |  |  |  |  |
| No (48) | 20.77 (5.47) |  | ref | 0.12 |
| Yes (24) | 17.96 (4.26) | 0.03 | -2.06 (-4.70, 0.58) |  |
| Social anxiety disorder |  |  |  |  |
| No (54) | 20.24 (5.02) |  | ref | 0.33 |
| Yes (18) | 18.61 (5.84) | 0.26 | -1.39 (-4.21, 1.43) |  |

N=number; SD=standard deviation; CI=confidence interval; ref=reference value of 1. <sup>‡</sup>Final models adjusted for child's age, sex and father's highest education level attained. <sup>∞</sup> [adjusted linear regression model constant=36.05, R<sup>2</sup>=0.21, F-ratio (5, 66) =3.46, p=0.008, N=72]. <sup>a</sup>based on Asthma Control Test (ACT) and childhood Asthma Control Test (cACT); Scores ranged from 8-34, low scores indicating poor asthma control and high scores indicating well controlled asthma.

### Risk factors associated with anxiety disorders

In this analysis the main outcome was any of the three major anxiety disorders among adolescents. We found that adolescents with anxiety disorders, compared to their counterparts without, were more likely to have been born in an urban area (than rural; AOR 3.42, 95% CI 1.29-9.09); raised in an urban area for their first five years of life (AOR 2.87, 95% CI 1.07-7.66); have fathers with tertiary education (compared to secondary or less; AOR 2.09, 95% CI 1.00-4.37); to have a father with a reported history of asthma (AOR 6.52, 95% CI 1.59-26.76); to report a history of other allergy-related diseases (rhinitis, allergic conjunctivitis or eczema; AOR 4.64, 95% CI 1.66-13.00); but there was no difference in allergic sensitisation (SPT to any of seven allergens; AOR 1.31, 95% CI 0.65-2.67), **Table 3**. These risk factors were similar to the asthma risk factors we reported from the parent case-control study^10^.

**Table 3:**
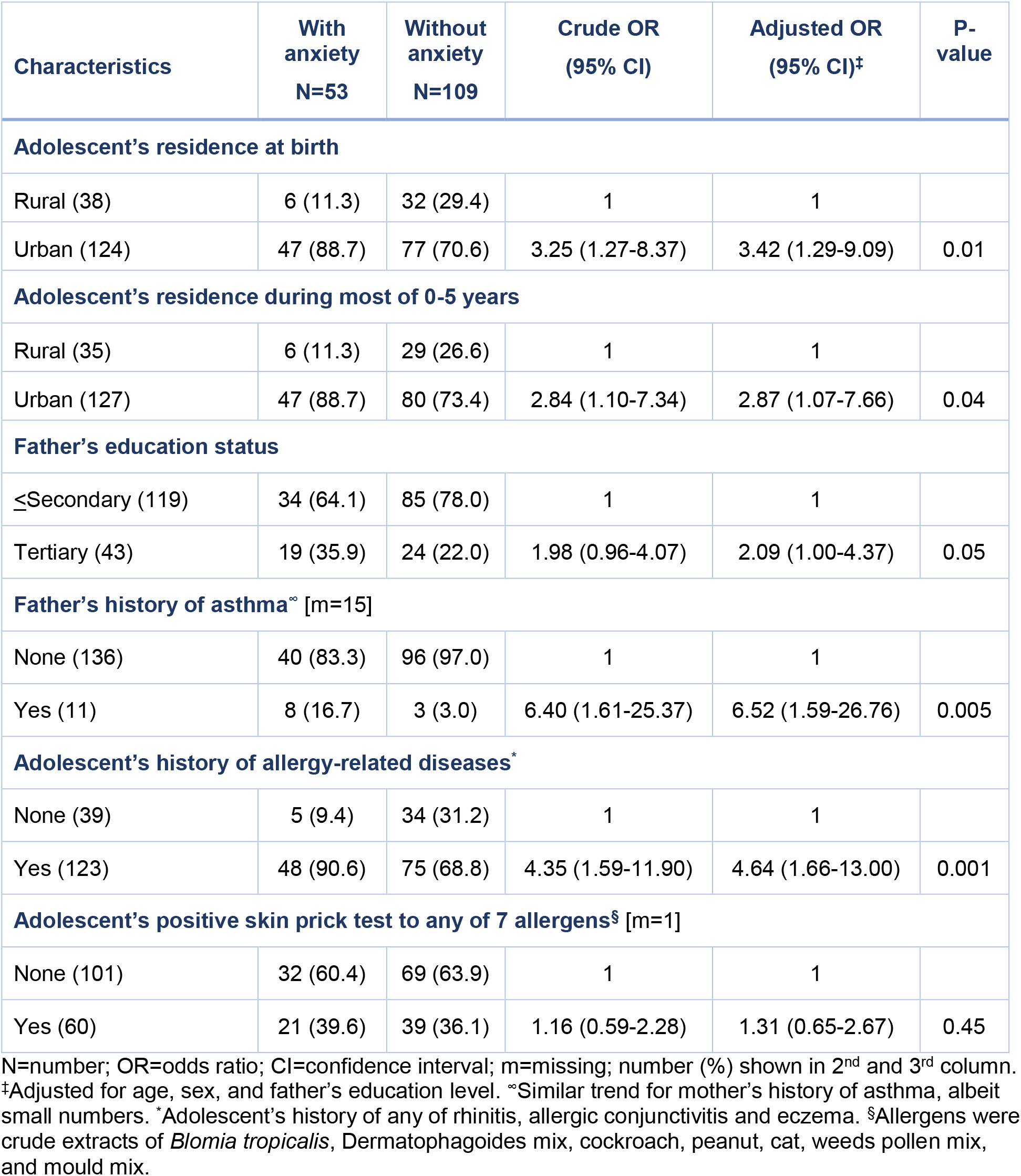
Risk factors associated with anxiety disorders among adolescents in Uganda (N=162)

Given the observed importance of area of residence in early life, father’s education status and adolescent’s asthma as risk factors for anxiety disorders, we explored how the different combinations of risk factors were associated with anxiety disorders. Firstly, compared to adolescents born in the rural areas and whose fathers had only primary or secondary education (reference group), adolescents born in urban areas and whose fathers had a tertiary education had the highest risk of anxiety disorders (AOR 8.67, 95% CI 2.14-35.10), **Table 4**.

**Table 4:** The association between the combined effects of residence in early life and father’s education with on anxiety disorders among adolescents in urban Uganda (N=162)

| Residence at birth | Father's education level | Anxiety disorder among adolescents |  | Crude OR (95% CI) | Adjusted OR (95% CI)<sup>‡</sup> | P-value |
| --- | --- | --- | --- | --- | --- | --- |
|  |  | Yes(n=53) | No(n=109) |  |  |  |
| Rural | Pri/sec | 3 (5.7) | 27 (24.8) | 1 | 1 |  |
| Urban | Pri/sec | 31 (58.4) | 58 (53.2) | 4.81 (1.35-17.13) | 1.02 (1.39-18.14) |  |
| Rural | Tertiary | 3 (5.7) | 5 (4.6) | 5.40 (0.84-34.80) | 5.34 (0.81-35.33) |  |
| Urban | Tertiary | 16 (30.2) | 19 (17.4) | 7.58 (1.93-29.70) | 8.67 (2.14-35.10) | 0.007 |

Pri=primary; sec=Secondary; n=number; OR=odds ratio; CI=confidence interval. 3<sup>rd</sup> and 4<sup>th</sup> columns show numbers (percentages). <sup>‡</sup>Adjusted for child's age and sex. Similar observations were made for adolescent's residence in the first five years of life.

Secondly, compared to adolescents born in the rural area and with no asthma (reference group), adolescents born in urban areas and with asthma had the highest risk for anxiety disorders (AOR 6.03, 95% CI 1.81-20.02), **Table 5**.

**Table 5:** Combined effects of residence in early life and current asthma as risk factors for anxiety disorders among adolescents in urban Uganda (N=162)

| Residence at birth | Asthma among adolescent | Anxiety disorders among adolescents |  | Crude OR (95% CI) | Adjusted OR (95% CI)<sup>‡</sup> |
| --- | --- | --- | --- | --- | --- |
|  |  | Yes (n=53) | No (n=109) |  |  |
| Rural | - | 4 (7.5) | 24 (22.0) | 1 | 1 |
| Urban | - | 2 (3.8) | 8 (7.3) | 1.50 (0.23-9.80) | 1.25 (0.18-8.76) |
| Rural | + | 15 (28.3) | 46 (42.2) | 1.96 (0.58-6.55) | 1.99 (0.58-6.84) |
| Urban | + | 32 (60.4) | 31 (28.5) | 6.19 (1.93-19.92) | 6.03 (1.81-20.02) |
| P-value=0.002 |  |  |  |  |  |

N=number; OR=odds ratio; CI=confidence interval; “-” refers to adolescent has no anxiety disorders, “+” refers to adolescent has an anxiety disorder. number (%) in 3<sup>rd</sup> and 4<sup>th</sup> column. <sup>‡</sup>Adjusted for child’s age, sex, and father’s education level. Similar observations were made for adolescent’s residence in the first five years of life.

In order to investigate further the hypothesis that both anxiety disorders and asthma may be due to shared early-life exposures, we additionally adjusted for urban residence at birth in the regression model assessing the association between anxiety disorders and asthma. We saw the newly adjusted odds ratio go towards the null value; compared to values in Table 1, the newly adjusted odds ratios reduced to 2.68 (95% CI 1.30-5.53) for any of three anxiety disorders, 4.49 (95% CI 1.48-13.56) for GAD, 5.43 (95% CI 2.11-14.02) for panic disorder, and 1.46 (95% CI 0.63-3.37) for social anxiety disorder. Thus, providing support for the “shared exposure” hypothesis.

### Psychological triggers for asthma and residence in early life

Given the positive association between anxiety disorders and urban residence in early life, we conducted an exploratory analysis to investigate the association between reporting psychological triggers for asthma and area of residence in early life, among all adolescents with asthma enrolled in the parent case-control study (N=275). We found a positive trend towards adolescents who reported psychological triggers for asthma being more likely to have been born in the city than adolescents without reported psychological triggers for asthma, a trend that was statistically significant for adolescents who spent most of their first five years of life in a town (AOR 2.28, 95% CI 0.88-5.86)) or the city (AOR 3.53, 95% CI 1.12-11.18); test for trend p=0.03, **Table 6**.

**Table 6:** Area of residence in early life and reported psychological triggers for asthma symptoms among adolescents with asthma in urban Uganda (N=275)

| Residence | Reported psychological trigger |  |  |  |
| --- | --- | --- | --- | --- |
|  | Yes (N=69) | No (N=206) | Adj. OR (95% CI)<sup>‡</sup> | P-value |
| Adolescent's residence at birth |  |  |  |  |
| Rural (48) | 10 (20.8) | 38 (79.2) | 1 | 0.17 |
| Town (186) | 43 (23.1) | 143 (76.9) | 1.07 (0.49-2.36) |  |
| City (41) | 16 (39.0) | 25 (61.0) | 2.14 (0.82-5.61) |  |
| Test for trend |  |  | P-value=0.11 |  |
| Adolescent's residence most of 0-5 years |  |  |  |  |
| Rural (42) | 6 (14.3) | 36 (85.7) | 1 | 0.08 |
| Town (200) | 51 (25.5) | 149 (74.5) | 2.28 (0.88-5.86) |  |
| City (33) | 12 (36.4) | 21 (63.6) | 3.53 (1.12-11.18) |  |
| Test for trend |  |  | P-value=0.03 |  |

Adj. OR=adjusted odds ratio; CI=confidence interval. 2<sup>nd</sup> and 3<sup>rd</sup> columns show numbers (percentages).
<sup>‡</sup>adjusted for child's age, sex and fathers' highest education level

## Discussion

We observed a positive association between anxiety disorders and asthma among adolescents in urban Uganda. We also found that adolescents with both asthma and anxiety disorders had had poorer asthma control than asthmatics without anxiety disorders. Additionally, early life risk factors associated with anxiety disorders among adolescents were similar to risk factors for asthma that we described previously, including urban residence in early life (compared to rural), parental tertiary education (compared to secondary or lower education), and adolescents’ own history of other allergy-related diseases^10^. The positive associations between anxiety disorders and asthma among adolescents were of similar magnitude to those previously reported from high-income countries^12, 13^.

Although the temporal relationship between anxiety disorders and asthma is not yet clearly understood, this study has demonstrated that both conditions share risk factors in early life. Of particular interest was the observation that the risk of anxiety disorders among adolescents in urban Uganda was related to their residence in early life, with the highest risk among adolescents born and raised in the urban areas. This is similar to what we described for asthma previously: compared to schoolchildren in urban Uganda who were born and raised rural areas, the risk of asthma was double among adolescents born and raised in any town and triple among the city born and raised^10^. Indeed, the highest risk for both conditions seems to be among schoolchildren born and raised in urban area whose parent had a tertiary education. These results suggest that the positive association between anxiety disorders and asthma during adolescence may have its roots in early life, perhaps due to shared environmental risk factors (and possibly underlying causes) in urban areas, different from exposures in rural areas. This hypothesis of shared environmental factors in early life is also supported by twin studies from Europe which have shown familial aggregation of both anxiety disorders and asthma among children, which could not be explained by genetic factors^16^.

Several studies in LMICs have consistently found a higher prevalence of asthma among children and adults in urban than rural areas, but the specific causative environmental exposures have not been identified^9^. It is likely that several environmental factors are at play, and initial ecological studies to decipher these have reported that lifestyle and socioeconomic factors have stronger overall effects on asthma prevalence than infrastructure factors^9, 25^. Indeed, there is increasing evidence for adverse effects of exposure to psychosocial factors in early life on increased risk of both asthma and anxiety disorders in later childhood: several studies have found that maternal anxiety disorders and parenting difficulties were positively associated with anxiety disorders in childhood^11, 26, 27^, and that maternal prenatal and postnatal psychosocial distress, and parenting difficulties were associated with increased risk of asthma (and allergy-related diseases) in childhood^11, 28-31^. Most studies investigating the adverse role of parental psychosocial distress on childhood asthma have been conducted in high-income countries, with only one study from sub-Saharan Africa that was conducted in South Africa^32^. It is important to investigate whether psychosocial distress or parenting difficulties are higher among urban dwellers than their rural counterparts in LMICs, and whether this could possibly contribute to the observed increased prevalence of asthma in urban areas. Such information would be vital in informing the design of intervention studies for the primary and secondary prevention of both anxiety disorders and asthma among children.

The observation of a positive association between anxiety disorders and poor asthma control has been made elsewhere^33, 34^. Since there was no follow-up in this study, temporality cannot be assumed. Nonetheless, this and the observation that a quarter of adolescents with asthma reported their symptoms were triggered by psychological factors (such as fear, anxiety, sadness, or excitement), emphasises the need to manage the two conditions concurrently, in order to improve asthma control and to minimise the frequency of asthma attacks. This may include the routine screening of asthmatics for anxiety disorders and the proper management of the latter^*35*^.

The parent study in which we nested this study was designed as an asthma prevalence case-control study, and therefore asthmatics were overrepresented compared to a general population. Therefore, we cannot estimate the population prevalence of anxiety disorders. The first study limitation was a lack of data on current residence (although adolescents were attending schools in urban Uganda, it is possible the rural-born adolescents return to rural areas during the school holiday and urban-born return to urban areas). The second limitation was the relatively small sample size which meant that we had reduced power for some analyses. Nevertheless, within the relatively small group of adolescents, we have been able to contribute data from Africa on the co-morbidity between anxiety disorders and asthma among adolescents. We also make the intriguing observation of a positive trend towards adolescents who reported psychological distress/excitement as a trigger for asthma being more likely to have been raised in the urban than rural areas. Are psychological triggers for asthma symptoms a manifestation of anxiety-related responses to learned threats that may be more prevalent in urban that rural areas? More studies will expand our understanding of the relationship between anxiety disorders and asthma.

## Conclusion

We confirm a positive association between asthma and anxiety disorders among adolescents in urban Uganda. In addition, we demonstrate a close similarity in early life risk factors associated with both anxiety disorders and with asthma among adolescents, suggesting shared underlying causes. This also implies that looking closely at the causes of anxiety disorders will provide more insight into the causes of asthma. More research is required to identify the adverse environmental factors in the urban areas in LMICs that increase both anxiety disorders and asthma, identify underlying mechanisms, and inform intervention studies for the prevention.

## Data Availability

Data will be made available on the London School of Hygiene and Tropical Medicine Data Compass.

## Acknowledgements

We extend our gratitude to the large team of researchers, at the MRC/UVRI and LSHTM Uganda research Unit, involved in the data collection and management for this study. We thank the study participants, their parents, the school and education authorities for their enthusiastic participation and contribution towards the success of this study.

